# Socioeconomic status, lifestyle, and hypertension prevalence of the older adults in China

**DOI:** 10.1101/2022.06.16.22276490

**Authors:** Jun Li, Meng Wei, Xiao Han, Hongman Wang

## Abstract

**Aims:** To examine the relationship between socioeconomic status (SES) and hypertension prevalence among the older adults in China.

**Method:** We used the panel data of the three rounds (2006, 2009, and 2011) of the China Nutrition and Health Survey to examine the relationship adjusted for lifestyle factors by multi-level logistic regression.

**Results:** Income and urbanicity levels are associated with hypertension in men. Men with low and middle income and living in low and middle urbanicity areas are more likely to have hypertension. Income and education are associated with hypertension in women. Women with low or middle income are more likely to have hypertension; women with lower education levels are less likely to have hypertension; women who never attended school are nearly as prone to get the disease as those who had high school or above education.

**Conclusion:** Income, and community urbanicity level are reversely correlated with hypertension prevalence among older Chinese men and women; hypertension prevalence rates of older Chinese women vary by education levels without a gradient.

---

With 11.4% of the Chinese population aged over 60 years old^1^ and an increased old-age dependency ratio caused by lower birth rates, the social care and health care of the older adults in the society had attracted great attention in China. In most middle-income countries, where the situation is very similar, hypertension ranks high among the non-communicable diseases that are replacing communicable diseases as the main causes of mortality and hypertension has a high rate of prevalence and low rates of awareness, treatment, and control in the older population.^2-4^

Previous studies have explored the relationship between SES and health in the general population and the older adult population in some regions in China.^5^ Health disparity by SES among this population has been reported.^6-12^ However, the relationship between hypertension and SES in this population in nationally representative samples remains unclear. Previous research also contains potential inaccuracy in the construction of some lifestyle factors, for example, using work-related PA as a proxy of overall PA.^13^

We used nationally representative longitudinal panel survey data to examined the relationship, controlling for lifestyle factors pinpointed by the World Health Organization for non-communicable diseases control—alcohol consumption, tobacco smoking, physical activity (PA, in MET-hours), total energy intake (TEI), and proportion of fat in total energy intake (as proportion in total calories, FIT) and demographic variables. The research question is what are the effects of the SES factors on hypertension in older men and older women in China?

We used the data of the 2006, 2009, and 2011 rounds of the China Nutrition and Health Surveys (CNHS, project website: http://www.cpc.unc.edu/projects/china), which had been conducted for 10 rounds between 1989 and 2015. The project investigated more than 7,200 urban and rural households and more than 30,000 individuals in 15 provinces chosen by multi-stage randomized sampling. We analyzed the 9,495 subjects aged 60 and over in the 9 provinces (Liaoning, Heilongjiang, Jiangsu, Shandong, Henan, Hubei, Hunan, Guangxi, Guizhou) covered by the CHNS in the three rounds. There were 2,876 subjects, 3,193 subjects and 3,426 subjects eligible in the three rounds, respectively. The total repeated measure rate of subjects (survey for at least two rounds) is 52.6% (82.34% in the 2006 and 2009 rounds, and 41.27% in the 2009 and 2011 rounds).

We represent subject SES by their annual per capita household income (PCHI), education level, and community urbanization index (CUI). Lifestyle factors controlled were smoking status, drinking status, TEI, FIT, and PA.

## Education

4 groups (1=Never attended school; 2=Primary school; 3=Junior middle school; 4=High school or above).

## PCHI groups

Three groups divided by the tertile cut-off points, with income adjusted against the 2011 Consumer Price Index.

## CUI groups

Three groups by tertile cut-off points of CUI, a composite urbanicity scale of 12 aspects, each scoring between 0 and 10.^14^

## Smoking status

yes or no, reported by subjects when the answer to the question of “whether there is a history of smoking.”

## Drinking status

yes or no, reported by subjects when the answer to the question of “whether you drank last year.”

## TEI

Three groups by tertile cut-off points. The average total energy intake per day was calculated against the food ingredient table. The diet data was obtained by 24 hours recall method.

## FIT groups

Three groups by tertile cut-off points. The proportion of fat in total energy intake= average calorie from fat per day/average total energy intake per day in the past three days (kcal). One gram of fat is calculated as 9 kcal of calories according to the method proposed by Xue.^15^

## PA groups

Three groups by tertile cut-off points. We used the “Compendium of Physical Activities 2011”^16^ to calculate weekly PA in metabolic equivalent values of the self-reported PAs (sum of “MET/hour of each activity” * “hours lasted for the activities”).

## Hypertension diagnosis

Subjects’ right arm blood pressure was measured three times in a sitting position after a 5-minute rest. The mean value of three measurements of the diastolic (and systolic) pressure was used as the diastolic (and systolic) pressure. Systolic blood pressure ≥140 mm Hg or diastolic blood pressure ≥90 mm Hg and/or taking antihypertensive drugs reported by the subjects is considered as having hypertension, according to the Chinese experts’ consensus for hypertension diagnosis and treatment in the older adults.^3^

The demographic control variables were age, ethnicity, and family size. For the selected characteristics of the subjects, see **Table 1**.

**Table 1.** Hypertension status and lifestyle of the subjects.

|  | Men |  |  |  |  |  | Women |  |  |  |  |  |
| --- | --- | --- | --- | --- | --- | --- | --- | --- | --- | --- | --- | --- |
|  | 2006 |  | 2009 |  | 2011 |  | 2006 |  | 2009 |  | 2011 |  |
|  | n | prpt (%) | n | prpt (%) | n | prpt (%) | n | prpt (%) | n | prpt (%) | n | prpt (%) |
| hypertensive | 369 | 33.21 | 571 | 50.49 | 610 | 45.97 | 419 | 32.38 | 662 | 52 | 735 | 48.77 |
| diagnosed | 991 | 89.76 | 972 | 75.35 | 1002 | 73.19 | 1104 | 87.2 | 1016 | 70.02 | 1070 | 68.37 |
| medicated | 76 | 36.36 | 261 | 83.92 | 301 | 82.69 | 117 | 43.17 | 378 | 87.3 | 423 | 85.98 |
| ever smoke | 760 | 64.35 | 761 | 58.99 | 807 | 58.95 | 158 | 11.57 | 108 | 7.44 | 129 | 8.24 |
| ever drank | 520 | 45.98 | 598 | 46.36 | 635 | 46.38 | 111 | 8.61 | 101 | 6.96 | 121 | 7.73 |
| FIT ≥ 30% | 567 | 50.44 | 668 | 51.98 | 760 | 56.05 | 669 | 52.51 | 816 | 56.86 | 885 | 57.92 |
|  | Mean | SD | Mean | SD | Mean | SD | Mean | SD | Mean | SD | Mean | SD |
| TEI | 2135.44 | 706.68 | 2143.7 | 660.88 | 2024.92 | 802.32 | 1811.43 | 601.2 | 1816.21 | 638.12 | 1728.47 | 709.97 |
| FIT | 29.62 | 11.97 | 31.18 | 11.03 | 31.99 | 11.42 | 30.68 | 12.1 | 32.12 | 11.41 | 33.19 | 12.21 |
| PAs | 67.22 | 116.46 | 82.05 | 132.13 | 92.07 | 132.92 | 96.87 | 109.57 | 86.04 | 102.32 | 90.88 | 108.62 |

Notes: n=number, propt=proportion, SD=standard deviation, diagnosed=had been previously diagnosed, FIT=fat in total energy intake, TEI=total energy intake, PAs= physical activities

The data in this study is nested and the between-group correlation coefficient values (men=0.74, women=0.75) and the design effect values (1108.78, 1250.5) indicated that a multi-level model is fit for use.^17-19^

We examined the relationship between SES and hypertension, after adjusting for the demographic and lifestyle factors, in multi-level logistic regression models (generalized linear models). One model for men and one model for women, respectively.

SPSS 23.0 was used for statistical analysis.

In the three rounds of the survey, 4,496 men (on average, 1,499 surveyed per round) and 5,000 women were surveyed (on average, 1,667 per round).

There is little gender difference in PCHI and CUI as subjects in the same households, regardless of gender, have the same PCHI, and subjects who reside in the same community have the same CUI value.

There is a vast difference between genders, and little difference between survey years in terms of education level, age, and family size. More women survived men and lived alone.

Women’s proportion of energy from fat is higher than that of men. More men had a history of smoking and reported drinking last year. Men’s daily cigarette consumption and drinking frequency were higher than women. Men’s smoking rate declined in the three rounds of the survey but was consistently higher than women (Table 1).

From 2006 to 2009, the prevalence of hypertension, the past diagnosis rate, and the rate of treatment in men and women increased (Table 1).

The accurate prediction value of the model increased with the addition of individual, family, community, and lifestyle variables until we obtained a satisfactory model for men and women respectively (Table 2). The model fit was judged based on the -2 log pseudo-fitness data. The smaller the value, the higher the model fit. The same applies for both modeling for men and women.

**Table 2.** Multi-level regression models.

|  | Men | Women |
| --- | --- | --- |
| **Fixed effect [OR (95% CI)]** |  |  |
| Intercept | 0.65 (0.34~1.21) | 1.20(0.44~3.20) |
| Age (Ref = $\geq 80$ years old) | | |
| 60-65 years old | 2.71*** (1.64~4.47) | 5.68*** (3.20~9.82) |
| 65-70 years old | 1.58 (0.96~2.61) | 4.26*** (2.50~7.25) |
| 70-75 years old | 1.36 (0.82~2.25) | 2.26*** (1.35~3.79) |
| 75-80 years old | 1.58 (0.95~2.12) | 0.80(0.49~1.30) |
| Education (Ref=high school) |  |  |
| Never attended school | 1.17 (0.75~1.82) | 0.94*(0.50~1.79) |
| Primary school | 0.97 (0.68~1.38) | 0.49*(0.26~0.89) |
| Junior middle school | 0.90 (0.62~1.31) | 0.50(0.26~0.97) |
| PCHI (Ref= high) |  |  |
| Low | 1.90*** (1.36~2.67) | 2.07*** (1.28~3.11) |
| Middle | 1.39* (1.02~1.90) | 1.96*** (1.33~2.90) |
| Family size (Ref= $\geq 2$ persons) | | |
| 1 person | 0.79 (0.48~1.30) | 0.78(0.50~1.20) |
| Urbanization level (Ref= high) |  |  |
| Low | 1.70*** (1.20~2.42) | 1.41(0.93~2.16) |
| Middle | 1.48* (1.07~2.04) | 1.00(0.68~1.44) |
| Smoking (Ref= Yes) | 0.77* (0.60~0.99) | 0.71(0.43~1.19) |
| Drinking (Ref= Yes) | 1.21 (0.96~1.53) | 0.68(0.42~1.13) |
| PA (Ref= high) |  |  |
| Low | 0.76 (0.56~1.04) | 0.88(0.60~1.29) |
| Middle | 0.89 (0.68~1.17) | 0.65** (0.47~0.90) |
| TEI (Ref = high) |  |  |
| Low | 1.11 (0.86~1.43) | 0.74(0.53~1.05) |
| Middle | 0.98 (0.76~1.25) | 0.65*(0.47~0.91) |
| FIT (Ref= high) |  |  |
| Low | 0.80 (0.61~1.04) | 1.68*** (1.25~2.25) |
| Middle | 0.85 (0.67~1.08) | 1.17(0.89~1.54) |
| **Randomized effect [variance (SE)]** |  |  |
| Intercept | redundant | 2.07*(0.16) |
| Age | 2.08*** (0.30) | 4.97(0.57) |
| Educational level | NS, O | NS, O |
| PCHI | 1.56*** (0.35) | 2.07*(0.16) |
| Family size | NS, O | NS, O |
| Urbanization | 1.54*** (0.32) | 4.97(0.57) |
| Drinking |  | 3.09*** (0.43) |
| PAs |  | 1.73*** (0.37) |
| Smoking | 1.08*** (0.30) | 3.649*** (0.33) |
| **Residual effect [variance (SE)]** |  |  |
| 2006 | 0.35*** (0.06) | 0.21*** (0.04) |
| 2009 | 0.51***(0.05) | 0.23***(0.03) |
| 2011 | 0.40***(0.05) | 0.27***(0.03) |
| **Summary** |  |  |
| Accurate prediction rate | 96.00% | 98.70% |
| -2 log pseudo-fitting | 16289 | 20384 |

Note: Ref: control group; \*= $p < 0.05$ , \*\*= $p < 0.01$ , \*\*\*= $p < 0.001$ ; NS, O=not significant, omitted; SE=standard error, PCHI=per capita household income; PA=physical activity; TEI=total energy intake; FIT=proportion of fat in total energy.

After adjusting for the demographic and lifestyle factors, men with low income and middle income (compared with high income, OR= 1.90(1.36∼2.67), *p* < 0.001 and 1.39(1.02∼1.90), *p*<0.05) and with low and middle CUI (compared with high CUI, OR= 1.70(1.20∼2.42), *p* < 0.001, and 1.48(1.07∼2.04), *p* < 0.05) are more likely to have the disease (Table 2).

For women, we found the association between income (PCHI) and hypertension (OR= 2.07***(1.28∼3.11), *p* < 0.001 for low income, and 1.96***(1.33∼2.90), *p* < for middle income) and the association between education level. Women with a lower education (primary schools) level are less likely to have hypertension (OR= 0.49(0.26∼0.89), *p* < 0.05). But it is worth noting that women who never attended school, very probably illiterate, are almost equal probability to have hypertension as those who had high school or above education (OR=0.94(0.50∼1.79, *p*<0.05) (Table 2).

We found that SES factors—income, urbanicity, and education, are related to hypertension, and that gender differences exist in the relation between SES factors and hypertension in the older adults in China.

Our study found that higher per capita household income functions as a protective factor of hypertension in the older adults. Similarly, Kaplan et al.^20^ had reported a significant inverse linear relationship between household income and the hypertension prevalence rate in the United States, but no evidence of such a relationship in Canada, and argued that social disparities and barriers to health care access and primary prevention in the United States may have played a role in it. We agree with Kaplan et al.’s explanation.^20^ As China had adopted a market-oriented healthcare system since its reform and opening-up at the end of the 1970s, income has become a much more important factor in obtaining living and healthcare resources. Many resources were obtained via a market mechanism, instead of rationing. Market-oriented policies stimulate efficiency in its economy, but at the cost of equity, as demonstrated here in health/disease disparity. To realize health equity, probably the strikingly severe income inequity as demonstrated in this study should be tackled first.

Our study confirms that people living in communities with higher urbanicity tend to have lower hypertension prevalence. Similar findings had been reported by previous studies. Buys et al.^21^ reported that the older adults with poorer community conditions have a higher prevalence of hypertension. Isaac and Laveist^22^ had reported that older people with convenience stores and recreational facilities have lower rates of hypertension, even if they live in communities of lower and medium SES. One possible mechanism is that with the decline of the mobility of older people, restrictions through neighborhood conditions became more prominent, rendering the older adults less capable of avoiding adverse socioeconomic conditions.^21^ Another possible mechanism might be that better income and living conditions facilitate the older adults to access quality preventive and curative health care and reduce anxiety and stress.^23^

Therefore, we argue that geriatric health management, especially the management of chronic diseases, depends on an array of conditions, including transportation, community health care services, built community environment. Future research could test such a hypothesis.

We found that women with a lower education (primary schools) level are less likely to have hypertension; women who never attended school are almost as likely to have hypertension as women who had high school or above education.

The findings of previous research on education level and hypertension in the older adults in China are inconsistent, possibly influenced by the age groups, locality, and ethnic groups studied. Lei et al.^24^ reported no gradient between the level of education and the prevalence of hypertension with a nationally representative sample data of the middle-aged and older adult population. Zuo et al.^25^ reported no gradient between education level and hypertension among the older adults of 12 counties in Hubei. However, in Xinjiang, Zhang et al.^26^ found a reversed correlation between education and hypertension among the older adults. Xinjiang is a northwestern region of China, where the proportion of Han Chinese is lower than that in other regions of China.

As for the possible mechanism underlying our finding, we speculate that older people with lower education have lower health literacy, which is essential in the management of their chronic diseases,^27^ and those with higher education tend to have lower physical activity level (which we had found in our study, would be published soon in another article), which could be a predictive factor of hypertension in the older adults in China. You et al.^28^ found that middle-aged and older adults in China who usually participated in the moderate-to-vigorous activity for more than 10 minutes were less likely to have hypertension and Miao et al.^29^ found similar results for the older adults in Beijing. More research is needed to verify the relationship between education and hypertension in the older adults.

The present study contributes to the literature by providing gender-specific results between SES and hypertension. It has been reported that the older adults with low SES in such countries as China^7,8,30,31^, Germany^32^, UK^33^, South Korea^34^, and the United States^20,35^, have worse health. However, previous studies ignore the gender difference in such a connection between SES and health. This study finds that the relationship between SES and hypertension in older men and women differ, which indicates that when considering the relation between social stratification and health of the older adults, gender should not be ignored.

We did not control the genetic variants, trace elements, diet structure, sleep, limitation of daily activities, cognitive abilities (which may affect their medication compliance and lifestyle), healthcare (such as medical security availability and access to medical services), SES of early life, pre-retirement occupation, and comorbidity in the study. The composite PA index that we use may obscure the effect difference of PA of high, medium, and low intensity. Future research could also consider dose, starting age and frequency of smoking and drinking and exposure to secondhand smoke, compare the accuracy of accelerometer-measured and self-recalled PA, and, if possible, use Mendelian randomization to examine the causality between SES and hypertension and the effect of lifestyle in the relationship.

Socioeconomic factors significantly correlate with hypertension in older men and women in China and that there are gender differences.

## Data Availability

The data used by this study is accessible at https://www.cpc.unc.edu/projects/china/data.

## Abbreviations

CNHS: China Nutrition and Health Surveys
CUI: Community Urbanization Index
FIT: proportion of fat in total energy intake
HHS: household size
MET-hours: metabolic equivalent hours OR odds ratio
PA: physical activity
PCHI: Per capita household income
SD: standard deviation
SE: standard error
SES: socioeconomic status
TEI: total energy intake

## Declaration

### 1. Ethics approval and consent to participate

This study used the data from the China Health and Nutrition Survey, which has obtained the ethics approval and subjects’ consent to participate. We consulted the ethics committee office at Peking University and was informed that no ethical review was required since we used a public database for extracting our data and no personal information was revealed in the article. Therefore, we ask for waiving of the ethics approval and consent to participate.

### 2. Consent for publication

Not applicable

### 5. Authors’ contributions

Jun Li: conceptualization, methodology, formal analysis, resources, data curation, and writing-original draft.

Meng Wei: conceptualization, methodology, formal analysis, resources; data curation; writing-original draft.

Xiao Han: writing-review & editing, submission.

Hongman Wang: conceptualization, methodology, resources, writing-review & editing, supervision, project administration, and funding acquisition.

All authors have read and approved the manuscript.

## 6. Acknowledgements

This research uses data from the China Health and Nutrition Survey (CHNS). We thank the National Institute of Nutrition and Food Safety, China Center for Disease Control and Prevention, Carolina Population Center, the University of North Carolina at Chapel Hill, the NIH (R01-HD30880, DK056350, and R01-HD38700) and the Fogarty International Center, NIH for financial support for the CHNS data collection and analysis of files from 1989 to 2006 and both parties plus the China-Japan Friendship Hospital, Ministry of Health for support for CHNS 2009 and future surveys. We thank Professor Yun Zhou at Department of Sociology, Peking University, and Dr. Yan Ning at China Health Education Center for their suggestions for the study.

## 7. Availability of data and materials

The data used by this study is accessible at https://www.cpc.unc.edu/projects/china/data.

